# Eosinophil and eosinophil-derived novel leukocyte ratios are strong predictors of the severity of acute coronary syndrome patients

**DOI:** 10.64898/2026.02.20.26346670

**Authors:** Chen Chen, Zai Hao Zhao, Lin Xu, Jia Ning Gao, Xiao Liu, Xiu Quan Quan, Yin Hua Zhang

## Abstract

Rapid prediction of the severity of acute coronary syndrome (ACS) is crucial for appropriate intervention in emergency department. Neutrophils (Neu), lymphocytes (Lym) and monocytes (Mon) and their ratios (Neu/Lym, NLR; Mon/Lym, MLR NeuxMon/Lym, SIRI) are acknowledged to be associated with the prediction of the severity and adverse outcome of ACS patients. Here, we analysed retrospectively eosinophils (Eos) and Eos-derived novel ratios (Neu/Eos, NER; Mon/Eos, MER; Neu x Mon/Eos, SIII; Neu/Eos x Lym, NEL; Mon/Eos x Lym, MEL; Neu x Mon/Eos x Lym, SV) of first admitted 1053 ACS patients within 24 hours of symptom onset to predict ST-segment elevation of myocardial infarction (STEMI), high Gensini score (H) and cardiac dysfunction (Killip Classification l to III grades). Results showed that Eos was significantly decreased in ST (n=227), Gensini (H) (n=311) and Killip I group (n=237) (P<0.05). All Eos-derived ratios (NER, MER, SIII, NEL, MEL, SV) were significantly higher with diagnostic severity (ST, Gensini (H), and Killip I group (P<0.05). ROC analysis revealed that SIII and SV predicted ST and Gensini (H) with high specificity and sensitivity, which were similar to that of NLR, MLR and SIRI. Conclusion: Eos and Eos-derived ratios, SIII and SV in particular, are strongly linked to the prediction of the severity of ACS, along with those of well-established leukocyte ratios. The new ratios of Eos hold significant importance in emergency department for quick evaluation of ACS patients.

## Introduction

Cardiovascular diseases (CVD) are the leading causes of death worldwide, with an estimated 19 million CVD-related deaths globally in 2020, representing 18.71% increase from 2010 ^1^. Acute coronary syndrome (ACS), particularly myocardial infarction (MI), is a major cause of fatal heart attack and mortality. During ACS, ischemia and necrosis occur in cardiac tissue, damaged myocardial cells, endothelial cells, and activated platelets release large amounts of inflammatory mediators and neurohormones. These signals are transported through the bloodstream to the bone marrow, stimulating hematopoietic stem cells and progenitor cells to the transition from a quiescent state into an active cell cycle, prioritizing differentiation into myeloid cells. Progenitors of granulocytes and monocytes (Mon) proliferate rapidly, accelerate maturation, and predominantly generate and release innate immune cells. Neutrophil (Neu)-derived extracellular traps (NETs) directly damage the myocardium, occlude microvessels, and strongly recruit Mon. Mon differentiate into M1-type macrophages, which are hyperinflammatory, while M2-type macrophages promote healing ^2^. Concurrently, adaptive immune cells (Lym) are suppressed, leading to a myeloid-lymphoid imbalance. Protective regulatory T cells and B cells decrease, while pro-inflammatory T cells become relatively active, resulting in sustained injury.

Eosinophils (Eos), which constitute less than 5% of peripheral blood leukocytes, are multifunctional granulocytes. Increasing evidence suggests their involvement in cardiovascular diseases, with studies confirming their protective role in the heart following myocardial infarction (MI) ^3^. Clinical data and animal experiments have demonstrated a decline in blood Eos counts post-acute MI ^4–7^, further corroborated by the presence of Eos aggregation in the infarcted area of autopsy specimens ^8^. Eos are essential for acquiring anti-inflammatory macrophages, inflammation resolution, and scar formation. Their deficiency leads to reduced levels of anti-inflammatory cytokines (e.g., IL-4, IL-5, IL-13, IL-10), increased pro-inflammatory mediators, enhanced infiltration of Neu and macrophages, and impaired macrophage polarization. IL-4 therapy can reverse the adverse remodeling phenotype observed in Eos-deficient states.

Multiple studies have emphasized the role of total white blood cell count in patients with acute MI. White blood cells serve as primary mediators of inflammtion and play a critical role in the host’s defense against injury ^9,10^. Some of these studies investigated the association between Neu-to Lym ratio (NLR) and AMI. In the context of ACS prognosis, NLR serves as a hematological biomarker reflecting inflammation, oxidative stress, and endothelial damage. By indicating the balance between these two types of inflammatory cells, it provides a more accurate representation of the body’s inflammatory status compared to single-cell type measurements ^11^. As an easily obtainable inflammatory marker, NLR is utilized for risk assessment in high-risk patients ^12^. Elevated NLR levels are observed in ST-segment elevation myocardial infarction (STEMI) patients undergoing percutaneous coronary intervention (PCI) with severe coronary artery stenosis, and increased NLR effectively predicts the occurrence of major adverse cardiac events (MACE) and heart failure ^13^. NLR can serve as an early diagnostic indicator for cardiac function and myocardial injury as well as prognosis in AMI patients. The higher the NLR, the greater the cardiac dysfunction and myocardial injury in AMI patients ^14^. MLR (Mon/Lym Ratio) is independently correlated with the severity of coronary artery disease and more accurately reflects the extent of coronary artery lesions than NLR ^10^. Circulating Mons are high-risk factors for cardiovascular events ^15^ and play critical roles in endogenous inflammatory responses as key components of the innate immune system. SIRI (Systemic Inflammatory Response Index) is a comprehensive measure of peripheral Neu, Mon, and Lym counts, and ACS patients exhibit significantly elevated SIRI levels compared to stable coronary heart disease patients ^16^. A study analyzing patients with ACS treated with PCI demonstrated that NLR, MLR, and SIRI have prognostic value in ACS patients, with SIRI outperforming other inflammatory markers in predicting major adverse cardiac events (MACE) ^17^.

Therefore, utilizing simple clinical test results to predict the occurrence of fatal diseases holds significant clinical value, particularly in emergency settings where rapid and convenient assessment tools can substantially reduce decision-making time and secure precious treatment windows for patients. Currently, commonly used clinical methods for assessing the severity of ACS, such as coronary angiography, are accurate but limited by invasiveness and prolonged duration, making them inadequate for emergency rapid screening. Complete blood count (CBC), as a routine emergency test, offers convenient and cost-effective parameter acquisition. Inflammatory cell ratios derived from CBC (e.g., SIII and SV in this study) provide novel possibilities for rapid ACS assessment. This study aims to analyze the association between Eos and related novel ratios with ACS severity, validate their efficacy as rapid emergency predictors, and provide clinicians with more efficient disease assessment tools to facilitate early risk stratification and individualized treatment strategies for ACS patients.

## Methods

A total of 1,773 hospitalized patients with ACS who underwent primary PCI and completed revascularization were enrolled in this retrospective study (from January 2016 to December 2018 at the Affiliated Hospital of Yanbian University, Jilin Province, China). Among them, 83 cases of old myocardial infarction were excluded, 567 cases of other diseases (including infectious diseases, aseptic inflammation, rheumatic immune diseases, allergies, and organ dysfunction) were excluded, and 70 cases with missing CBC data were deleted, leaving 1,053 cases in the final cohort. Patients admitted to the hospital Emergency department were not involved in the designing of current research or the outcome measures, because this is a retrospective study. This study has been approved by the Institutional Review Board of Yanbian University (Yanbian University Ethics: 20250185). The study was conducted in accordance with the relevant principles of the Declaration of Helsinki.

Preoperative venous blood was collected from the patient, and general information (age, gender, BMI), medical history, heart rate, blood pressure, and CBC were obtained. The proportion of inflammatory cells was calculated based on the Neu, Mon, Lym, Eos in the CBC. The following ratios were derived: NLR = Neu/Lym, MLR = Mon/Lym, (Systemic Inflammatory Response Index, SIRI) = NeuxMon/Lym, NER = Neu/Eos, MER = Mon/Eos, SIII = NeuxMon/Eos, NEL = Neu/EosxLym, MEL = Mon/EosxLym, SV = NeuxMon/EosxLym.

All patients were assessed using the Gensini score based on coronary angiography (CAG) results. The Gensini score= sum of the stenosis severity scores × lesion location coefficients. 1. Stenosis severity score: Calculated based on the diameter of the most severe stenosis in the coronary artery. Stenosis severity: 0% (0 points), 1%–25% stenosis (1 point), 26%–50% stenosis (2 points), 51%–75% stenosis (4 points), 76%–90% stenosis (8 points), 91%–99% stenosis (16 points), 100% occlusion (32 points). 2. Lesion location coefficient: Different vessels have varying importance in myocardial blood supply, so corresponding coefficients are applied. Left main (coefficient 5), proximal left anterior descending or left circumflex (coefficient 2.5), mid-left anterior descending or left circumflex (coefficient 1.5), distal left anterior descending or left circumflex (coefficient 1), right coronary artery (coefficient 1), first diagonal or posterior descending (coefficient 1), second diagonal or posterior collateral or other small collateral branches (coefficient 0.5). Scoring criteria: normal (Gensini score = 0), low-risk group (1 ≤ Gensini score ≤ 20), intermediate-risk group (21 ≤ Gensini score ≤ 39), high-risk group (40 ≤ Gensini score ≤ 99).

Cardiac function: Killip class I, no signs of heart failure, but PCWP (pulmonary capillary wedge pressure) may be elevated, with a mortality rate of 0-5%. Class II, mild to moderate heart failure, with pulmonary rales present in less than 50% of the lung fields, possible third heart sound gallop rhythm, persistent sinus tachycardia or other arrhythmias, elevated venous pressure, and radiographic evidence of pulmonary congestion, with a mortality rate of 10-20%. Class III, severe heart failure, with acute pulmonary edema, pulmonary rales present in more than 50% of the lung fields, and a mortality rate of 35-40%. Class IV, cardiogenic shock, with systolic blood pressure <90 mmHg, urine output <20 mL/h, cold and clammy skin, cyanosis, tachypnea, and pulse rate>100 beats/min, with a mortality rate of 85-95%. Cardiac function assessment is based on the diagnostic results of the attending physician.

Statistical Analysis Data analysis was performed using SPSS 23.0 (IBM) statistical software. To assess the overall distribution characteristics of data, descriptive analysis of skewed data was conducted using quartiles (M (P25, P75)). Nonparametric statistical analysis was employed for intergroup comparisons. The receiver operating characteristic (ROC) curve was used to investigate the relationship between the proportion of inflammatory cells and the ECG diagnosis of ACS, while the Gensini score was used to analyze the relationship between vascular stenosis severity and cardiac function classification. A P-value <0.05 was considered statistically significant.

## Results

A total of 1,053 patients with ACS were enrolled in this study, all of whom underwent PCI within 24 hours of symptom onset. Among them, 649 were male, accounting for 61.6%. The age quartile distribution was as follows: median age 61 years, lower quartile 54 years, and upper quartile 68 years. Basic demographic characteristics, medical history, and subgroup descriptions are presented in **Table 1**. The median systolic blood pressure was 130 mmHg, the median diastolic blood pressure was 80 mmHg, the median mean arterial pressure was 96 mmHg, and the median heart rate was 76 beats per minute. Smoking history was present in 45.7% of patients, diabetes in 20.1%, and hypertension in 57.1%.

**Table 1.** Description of Basic Information of 1053 Hospitalized Patients with Chest Pain.

| Parameter | Characteristic | ALL n=1053 |  |  |  |
| --- | --- | --- | --- | --- | --- |
| Demographics | Age (y) | 61 (54, 68) |  |  |  |
|  | Male (%) | 649 (61.6) |  |  |  |
|  | BMI (kg/m<sup>2</sup>) | 24.52 (22.19, 26.98) |  |  |  |
|  | SBPmmHg | 130 (120, 147) |  |  |  |
|  | DBPmmHg | 80 (70, 90) |  |  |  |
|  | MAPmmHg | 96 (90, 108) |  |  |  |
|  | Heart rate | 76 (68, 82) |  |  |  |
| Medical history, n (%) | Smoking | 482 (45.7) |  |  |  |
|  | Diabetes | 212 (20.1) |  |  |  |
|  | Hypertension | 602 (57.1) |  |  |  |
| Group (%) | Electrocardiogram | CON(12) | UA(48) | NST(18) | ST(22) |
|  | Gensini score | CON(18) | L(34) | M(18) | H(30) |
|  | (Killip)Cardiac function | CON(57) | I(23) | II(15) | III(6) |

The counts of inflammatory cells Neu and Mon were higher in the severe ACS group, with a more pronounced gradual increase in Neu (**Table 2A-C, Figures 1 Ai-Ci**). Lym and Eos exhibited opposite trends. The median values of Eos in the UA, NST, and ST groups were 0.12,0.08, and 0.06. In Gensini score of the L, M, and H groups, the median values were 0.11,0.09, and 0.07. A significant decrease in Eos was observed with increasing severity of ACS and more severe coronary artery stenosis (P<0.05). Notably, the changes in inflammatory cells starts from Killip I group to higher severity heart failure groups (**Table 2C, Figure 1Ci**). The median and quartile values of EOS in the heart function I, II, and III groups were 0.07 (0.03,0.13),0.07 (0.01,0.14), and 0.07 (0.02,0.18). Compared with the control group, I, II, or III/IV showed statistically significant differences (P<0.05). The results indicate that the Eos count gradually decreases with the severity of ACS which is similar to that of Lym, but Neu and Mon are increased.

**Figure 1.**
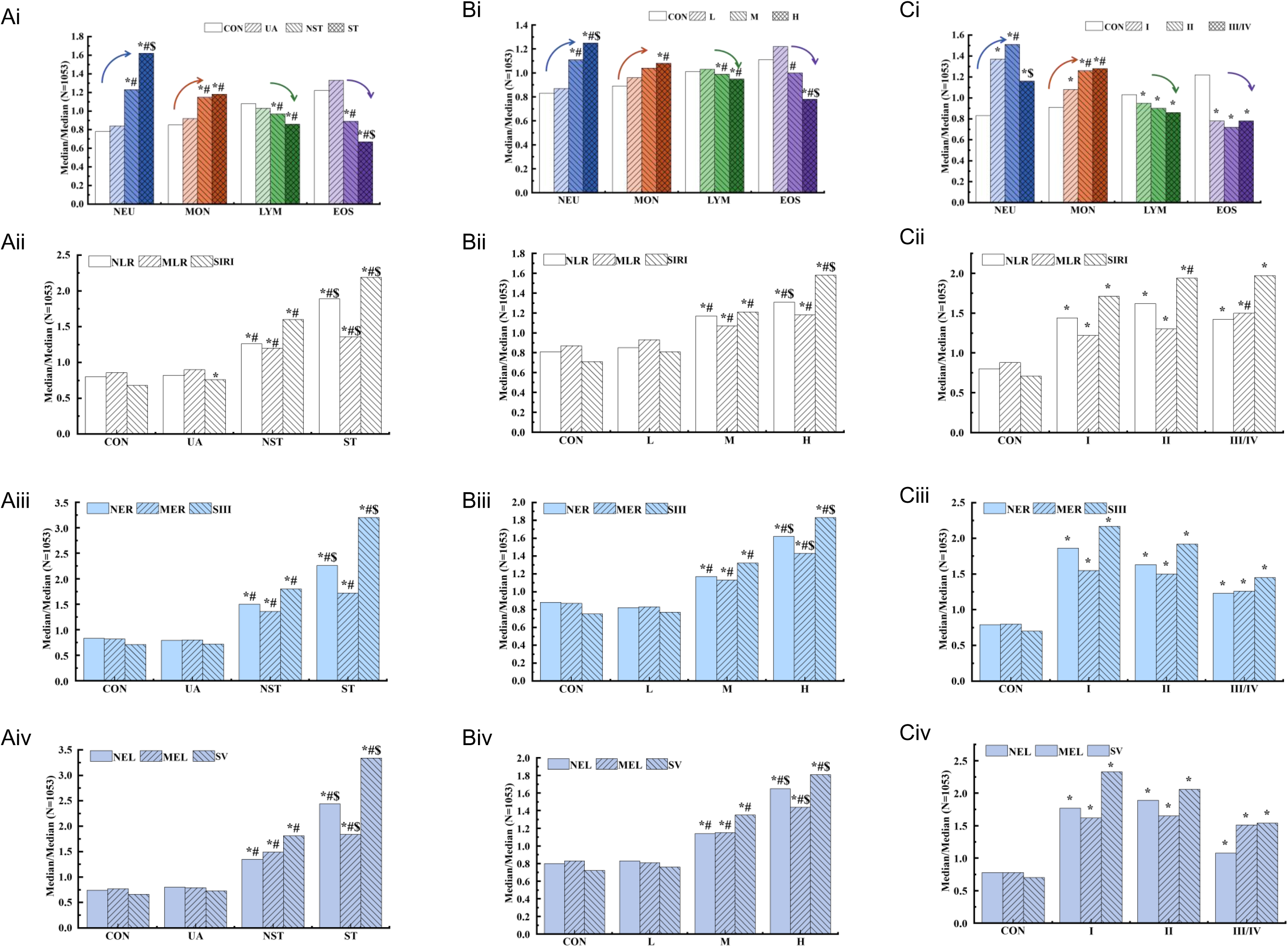
Inflammatory cells and Eos-derived novel leukocyte ratios in different groups of ACS patients. **A, B, C:** Median ratios of Neu, Mon, Lym, Eos to total cells and the ratios (NLR, MLR, SIRI, NER, MER, SIII, NEL, MEL, SV) in ACS patients grouped by Electrocardiology (**A**); Gensini score (**B**) and Killip grading (**C**). **i**. The median ratios of Neu, Mon, Lym, Eos to total cells (n=1053) in the Con, UA, NST, and ST grouped by ECG (**Ai**); in the Gensini score Con, L, M, and H groups (**Bi**); in the Con, I, II, and III/IV grouped by cardiac function (**Ci**). ii. The median ratios of NLR, MLR and SIRI in the Con, UA, NST, and ST grouped by ECG (**Aii**); in the Gensini score Con, L, M, and H groups (**Bii**); in the Con, I, II, and III/IV grouped by cardiac function (**Cii**). iii. The median ratios of NER, MER, SIII in the Con, UA, NST, and ST grouped by ECG (**Aiii**); in the Gensini score Con, L, M, and H groups (**Biii**); in the Con, I, II, and III/IV grouped by cardiac function (**Ciii**). iv. The median ratios of NEL, MEL, SV in the Con, UA, NST, and ST grouped by ECG (**Aiv**); in the Gensini score Con, L, M, and H groups (**Biv**); in the Con, I, II, and III/IV grouped by cardiac function (**Civ**).

**Table 2.** Descriptive Analysis of Inflammatory Cells in ACS Patients Grouped by ECG, GENSINI Score, and Cardiac Function.

|  |  |  |  |  |  |  |
| --- | --- | --- | --- | --- | --- | --- |
| A | Electrocardiogram | Parameter($10^9/L$) | CON (n=127) | UA (n=508) | NST (n=191) | ST (n=227) |
|  |  | NEU (1.8–6.3) | 3.35 (2.57, 4.41) | 3.59 (2.87, 4.66) | 5.28 (4.10, 7.57)<sup>*#</sup> | 6.95 (5.18, 9.52)<sup>*#</sup> |
|  |  | MON (0.1–0.6) | 0.45 (0.37, 0.61) | 0.49 (0.38, 0.6) | 0.61 (0.46, 0.79)<sup>*#</sup> | 0.63 (0.47, 0.86)<sup>*#</sup> |
|  |  | LYM(1.1–3.2) | 2.14 (1.75, 2.5) | 2.04 (1.68, 2.52) | 1.94 (1.41, 2.44)<sup>*#</sup> | 1.71 (1.26, 2.36)<sup>*#</sup> |
|  |  | EOS (0.02–0.5) | 0.11 (0.07, 0.18) | 0.12 (0.07, 0.21) | 0.08 (0.03, 0.14)<sup>*#</sup> | 0.06 (0.01, 0.13)<sup>*#</sup> |
| B | Gensini score | Parameter($10^9/L$) | CON (n=191) | L (n=358) | M (n=193) | H (n=311) |
|  |  | NEU (1.8–6.3) | 3.57 (2.64, 4.64) | 3.72 (2.94, 5) | 4.77 (3.53, 6.96)<sup>*#</sup> | 5.38 (4, 8.0)<sup>*#</sup> |
|  |  | MON (0.1–0.6) | 0.47 (0.38, 0.6) | 0.51 (0.39, 0.63) | 0.55 (0.43, 0.74)<sup>*#</sup> | 0.57 (0.44, 0.74)<sup>*#</sup> |
|  |  | LYM(1.1–3.2) | 2.01 (1.68, 2.44) | 2.04 (1.65, 2.57) | 1.98 (1.51, 2.44) | 1.89 (1.37, 2.44)<sup>*#</sup> |
|  |  | EOS (0.02–0.5) | 0.1 (0.06, 0.17) | 0.11 (0.07, 0.21) | 0.09 (0.05, 0.16)<sup>#</sup> | 0.07 (0.02, 0.15)<sup>*#</sup> |
| C | Cardiac function (Killip) | Parameter($10^9/L$) | CON (n=598) | I (n=237) | II (n=156) | III/IV (n=62) |
| | | NEU (1.8–6.3) | 3.55 (2.79, 4.54) | 5.88 (4.38, 8.11)<sup>*</sup> | 6.49 (4.74, 8.97)<sup>*#</sup> | 4.97 (3.76, 8.82)<sup>*\$</sup> |
|  |  | MON (0.1–0.6) | 0.48 (0.38, 0.59) | 0.57 (0.46, 0.76)<sup>*</sup> | 0.67 (0.48, 0.87)<sup>*#</sup> | 0.68 (0.48, 0.88)<sup>*#</sup> |
|  |  | LYM(1.1–3.2) | 2.05 (1.71, 2.51) | 1.90 (1.36, 2.5)<sup>*</sup> | 1.79 (1.36, 2.45)<sup>*</sup> | 1.71 (1.15, 2.19)<sup>*</sup> |
|  |  | EOS (0.02–0.5) | 0.11 (0.07, 0.21) | 0.07 (0.03, 0.13)<sup>*</sup> | 0.07 (0.01, 0.14)<sup>*</sup> | 0.07 (0.02, 0.18)<sup>*</sup> |

\* p value <0.05 compared with CON group, # p value <0.05 compared with UA, L, I group, \$ p value <0.05 compared with NST, M, II group.

Inflammatory cell ratios with Lym as the denominator was presented in **Table 3A and Figure 1Aii-Cii**. Results showed that the medians of NLR, MLR, and SIRI were the highest in the ST group compared to CON, UA, and NST (P <0.05). The descriptive analysis of inflammatory cell ratios with Eos as the denominator were shown in **Table 3B** and **Figure 1Aiii-Ciii**. Results showed that the medians of NER and SIII in the ST group were the highest than those in CON, UA, and NST (P <0.05). When Lym and Eos were combined as the denominator **(Table 3C, Figure 1Aiv-Civ)**, the medians of SV were the highest in the ST groups indicating high predictive value.

**Table 3.** Descriptive statistical analysis of and inflammatory cell ratios by electrocardiogram in the overall ACS data.

|  | Parameter | CON (n=127) | UA (n=508) | NST (n=191) | ST (n=227) |
| --- | --- | --- | --- | --- | --- |
| A | NLR | 1.69 (1.23, 2.17) | 1.74 (1.33, 2.35) | 2.68 (2.05, 4.5)<sup>*#</sup> | 4.01 (2.32, 7.04)<sup>*##</sup> |
|  | MLR | 0.22 (0.18, 0.29) | 0.23 (0.18, 0.31) | 0.31 (0.24, 0.42)<sup>*#</sup> | 0.35 (0.26, 0.47)<sup>*##</sup> |
|  | SIRI | 0.75 (0.53, 1.16) | 0.84 (0.57, 1.24)<sup>*</sup> | 1.77 (1.07, 2.58)<sup>*#</sup> | 2.42 (1.37, 4.09)<sup>*##</sup> |
| B | NER | 30.35 (16.52, 49.76) | 29.10 (16.72, 55.88) | 54.92 (29.54, 157.4)<sup>*#</sup> | 82.86 (36.33, 322.5)<sup>*##</sup> |
|  | MER | 4.11 (2.42, 7.33) | 4.00 (2.33, 7.17) | 6.80 (4.35, 12.91)<sup>*#</sup> | 8.60 (4.20, 25.0)<sup>*#</sup> |
|  | SIII | 13.96 (7.03, 24.29) | 13.97 (8.11, 27.69) | 35.22 (17.08, 83.65)<sup>*#</sup> | 62.50 (22.9, 201.24)<sup>*##</sup> |
| C | NEL | 13.29 (7.15, 25.94) | 14.27 (7.81, 28.77) | 24.12 (13.55, 101.5)<sup>*#</sup> | 43.69 (14.78, 196.1)<sup>*##</sup> |
|  | MEL | 1.88 (1.14, 3.27) | 1.92 (1.10, 3.66) | 3.61 (1.85, 7.36)<sup>*#</sup> | 4.47 (1.91, 14.91)<sup>*##</sup> |
|  | SV | 6.11 (3.66, 11.45) | 6.72 (3.81, 13.45) | 16.77 (7.53, 51.5)<sup>*#</sup> | 30.87 (10.19, 125.9)<sup>*##</sup> |

\* p value <0.05 compared with CON group, # p value <0.05 compared with UA group, \$ p value <0.05 compared with NST group.

In Gensini group, the medians of NLR and SIRI were significantly higher in Group H compared to Groups CON, L, and M (P <0.05) (**Table 4A, Figure 1Bii**). When Eos was the denominator (**Table 4B**) or when Lym and Eos were combined as the denominator (**Table 4C**), the inflammatory cell ratios were higher than those with Lym as the denominator. The median and quartiles of SIII and SV in Group H were significantly higher than those in Groups CON, L, and M (P <0.05). The results indicate that the more severe the ACS, the higher the Eos-related inflammatory cell ratios.(**Figures 1Bii-iii**).

**Table 4.** Descriptive statistical analysis of and inflammatory cell ratios by Gensini score in the overall ACS data.

|  | Parameter | CON (n=191) | L (n=358) | M (n=193) | H (n=311) |
| --- | --- | --- | --- | --- | --- |
| A | NLR | 1.72 (1.29, 2.41) | 1.80 (1.33, 2.47) | 2.48 (1.57, 4.03)<sup>*#</sup> | 2.79 (1.87, 5.31)<sup>*#</sup> |
|  | MLR | 0.23 (0.18, 0.32) | 0.24 (0.18, 0.32) | 0.28 (0.22, 0.39)<sup>*#</sup> | 0.31 (0.23, 0.41)<sup>*#</sup> |
|  | SIRI | 0.79 (0.56, 1.39) | 0.90 (0.58, 1.38) | 1.33 (0.85, 2.37)<sup>*#</sup> | 1.74 (0.97, 3.1)<sup>*#</sup> |
| B | NER | 32.17 (17.62, 55.27) | 29.86 (16.37, 55.8) | 42.86 (24.59, 100.83)<sup>*#</sup> | 59.4 (26.5, 193.88)<sup>*#</sup> |
|  | MER | 4.33 (2.43, 7.55) | 4.17 (2.38, 7.19) | 5.64 (3.14, 11.86)<sup>*#</sup> | 7.14 (3.63, 15.56)<sup>*#</sup> |
|  | SIII | 14.64 (7.81, 27.55) | 15.1 (8.15, 28.18) | 25.84 (11.31, 64.18)<sup>*#</sup> | 35.78 (15.03, 96.98)<sup>*#</sup> |
| C | NEL | 14.40 (8.19, 28.62) | 14.88 (7.76, 29.58) | 20.53 (10.73, 52.47)<sup>*#</sup> | 29.51 (13.08, 101.84)<sup>*#</sup> |
|  | MEL | 2.01 (1.19, 3.74) | 1.96 (1.11, 3.77) | 2.79 (1.36, 6.01)<sup>*#</sup> | 3.5 (1.74, 10.02)<sup>*#</sup> |
|  | SV | 6.66 (3.9, 13.95) | 7.05 (3.81, 13.63) | 12.47 (5.49, 32.79)<sup>*#</sup> | 16.72 (7.1, 63.22)<sup>*#</sup> |

\* p value <0.05 compared with CON group, # p value <0.05 compared with L group, \$ p value <0.05 compared with M group.

In cardiac function group (**Table 5**), the medians of NLR, MLR and SIRI were higher in from cardiac function classes I, but no significant differences were observed among classes I, II, and III/IV (p>0.05, **Table 5A, Figure 1Cii**). For EOS-related inflammatory cell ratios, the medians of SIII and SV in cardiac function class I were statistically higher compared to CON (P<0.05, **Table 5B,C, Figure 1Ciii-iv**). The results indicate that cardiac dysfunction is associated with elevated inflammatory ratios, particularly SIII and SV, but these ratios do not continue to increase with worsening cardiac function.

**Table 5.**
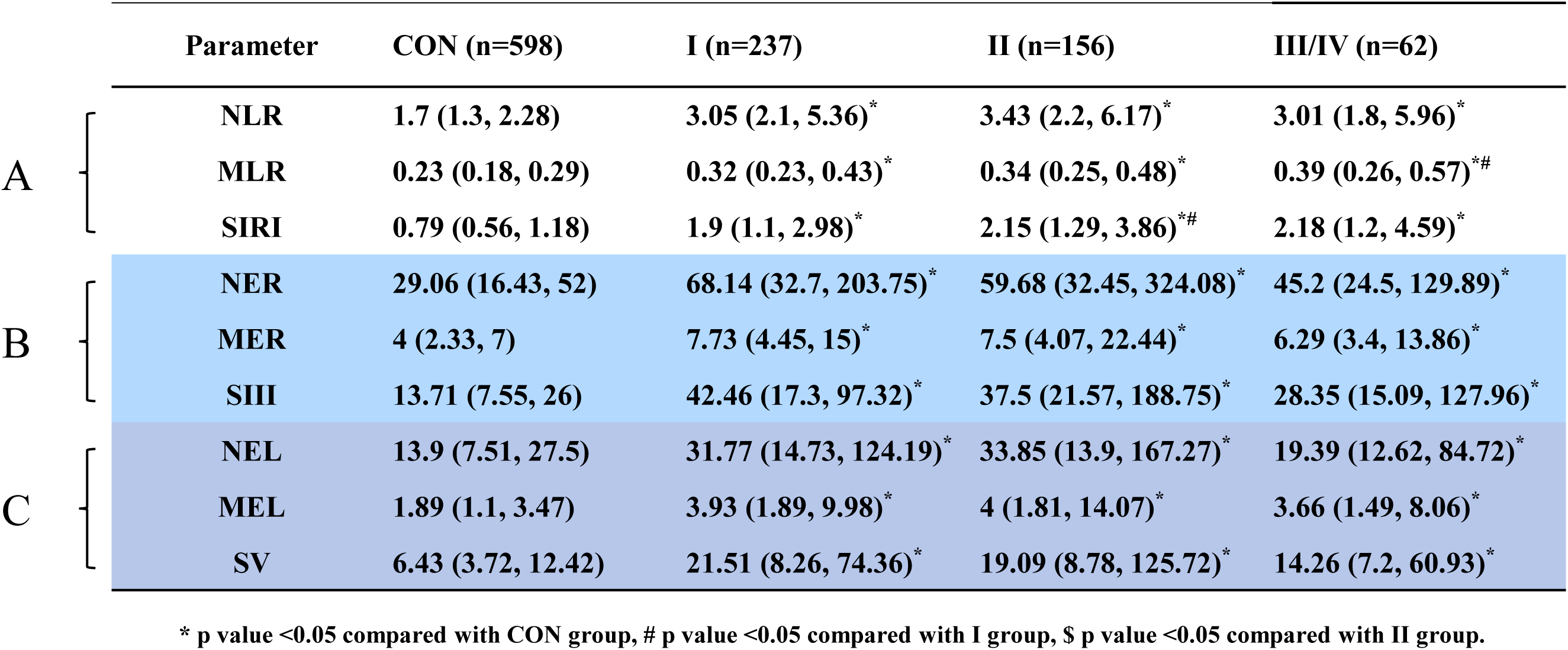
Descriptive statistical analysis of and inflammatory cell ratios by cardiac function in the overall ACS data.

The optimal cutoff values, sensitivity, and specificity for each group were determined using ROC curves (**Supplementary Table 1**). The cutoff value was selected based on the linear graph when the lesion was stable and the proportion of inflammatory cells showed significant changes (**Supplementary Figure 1**). As shown in **Figure 2**, the ROC curves for SIII and SV were comparable to NLR, MLR, SIRI in predicting ST, high-risk coronary artery stenosis, and cardiac function class I all exhibited an AUC value>0.7, indicating the validity for the disease diagnosis.

**Figure 2.**
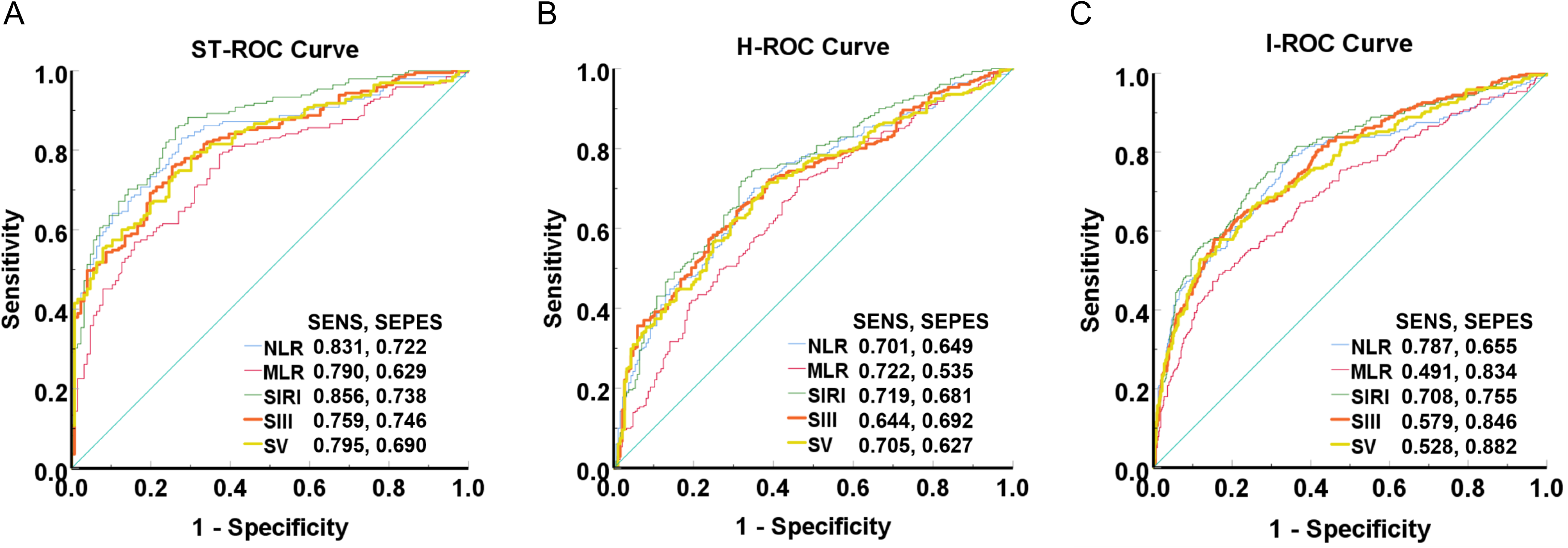
ROC curves of NLR, MLR, SIRI, SIII, and SV for predicting different severity levels of ACS, and the sensitivity and specificity of the inflammatory ratio. A. ROC curve of inflammatory ratio for predicting STEMI. B. ROC curve of inflammatory ratio for predicting high-risk coronary artery stenosis through Gensini score. C. ROC curve of inflammatory ratio for predicting cardiac function through Killip classification.

## Discussion

During the onset of ACS, the aggregation and changes of inflammatory cells in the bloodstream can predict the occurrence of future ischemic events. This study retrospectively analyzed data from 1,053 patients who underwent percutaneous coronary intervention (PCI) within 24 hours of symptom onset. In patients with ACS, we established new eosinophil-derived ratios (NER, MER, SIII, NEL, MEL, SV). The results demonstrated that early peripheral blood Eos was decreased with ACS severity, coronary stenosis, and cardiac dysfunction. All eosinophil-derived ratios (NER, MER, SIII, NEL, MEL, SV) were significantly elevated in groups with severe disease (ST-segment elevation, Gensini score (H)) and Killip class I (P<0.05), similar to NLR, MLR, and SIRI. Furthermore, eosinophil-derived ratios SIII and SV were confirmed to have high specificity and sensitivity for early prediction of ACS severity.

Immune dysregulation and hyperactivation-induced inflammatory cascades may mediate myocardial injury and are associated with adverse clinical outcomes in patients with ACS. Traditionally, CBC have been used as biomarkers to explore these associations, with particular attention paid to Neu, Mon, Lym counts. Recent studies have demonstrated that the WBC profile in patients with AMI is correlated with prognosis ^18^. Neu are among the earliest cells recruited from the bloodstream following cardiac ischemia, while Mon counts are associated with expanded infarct size and impaired contractile function ^19^. Decreased Lym counts are linked to an increased risk of MI ^20^. Our findings revealed a progressive upward trend in Neu and Mon counts in the CON, UA, NST, and ST groups or in the CON, L, M, and H groups. Lym counts were decreased with increasing disease severity in the ECG-based grouping. In the Gensini scoring system, Lym counts was decreased with increasing bifurcation in the vascular stenosis group.

It is noteworthy that Eos counts vary in cardiovascular disease risk prediction studies. Jiang et al. found that peripheral Eos counts were lowest in patients with the largest AMI area. They observed that patients with troponin I ≥20 ng/mL had significantly lower Eos counts (ELR) compared to those with troponin I <20 ng/mL ^21^. Previous studies have demonstrated that Eos participate in intravascular thrombus formation through their enzymatic-dependent production of strong endogenous thrombin ^22^. Eos play a critical role in thrombus formation, progression, and rupture ^6^, and contribute to coronary artery occlusion by promoting thrombus growth ^23^. Histological analysis of tissue samples obtained from thrombus aspiration therapy in ACS patients revealed extensive Eos infiltration within the thrombus ^23^. In our results, Eos counts decreased with disease severity according to ECG-diagnosed groups, with medians and quartiles in the UA, NST, and ST groups being (0.12 (0.07,0.21),0.08 (0.03,0.14),0.06 (0.01,0.13), respectively). When overall data were stratified by Gensini score, the medians and quartiles in the L, M, and H groups were (0.11 (0.07,0.21),0.09 (0.05,0.16),0.07 (0.02,0.15). Lower Eos counts were associated with more severe CAD. We also observed significantly lower Eos counts in the NST and ST groups compared to the CON and UA groups. In STEMI patients undergoing PCI, reduced EOS% was associated with in-hospital MACEs. Low peripheral Eos counts predict an increased risk of severe CAD, STEMI, or NSTEMI ^24^.

We cautiously hypothesize that the reduction in peripheral circulating Eos may be induced by extensive thrombosis; however, the underlying mechanisms require further investigation. Elevated cortisol levels due to acute stress response during acute MI may also contribute to decreased peripheral Eos counts ^25^. Conversely, significantly elevated peripheral Eos levels in patients with ACS independently predict the severity of CAD ^26^. As Jiang suggested, circulating Eos reflect the extent of MI. Toor et al. found that increased Eos counts were associated with elevated long-term follow-up mortality risk, making them useful biomarkers for risk stratification and mortality prediction in CAD patients. These clinical studies demonstrate that Eos counts are associated with cardiovascular disease risk. In long-term follow-up, ACS patients who experienced adverse cardiovascular events exhibited higher Eos levels ^18,27,28^, while low Eos levels within a short period (e.g., 24 hours) significantly increased the risk of adverse cardiovascular events ^29^. This may be attributed to factors such as disease type, cohort differences, data collection methods, analysis timing, and study duration.

Extensive clinical data indicate that the NLR in peripheral blood can effectively predict the risk of death or major adverse cardiac events in patients with ACS undergoing PCI ^30^. These cellular subsets may serve as important laboratory markers for prognosis and clinical management in such patients. Studies have also demonstrated that in NSTE-ACS, NLR is associated with the SYNTAX score ^31^. Furthermore, prior research has shown that both neutrophilia and lymphopenia are independently and significantly correlated with increased risks of post-AMI complications and mortality ^32–34^. Numerous studies have investigated the predictive value of NLR for adverse outcomes and mortality after myocardial infarction, with higher NLR values exhibiting stronger predictive power ^35–37^. Oncel R.C. et al. examined the impact of NLR and the Grace risk score on in-hospital cardiac events in patients with STEMI. The results revealed that a higher NLR at admission was associated with increased rates of in-hospital cardiac death, recurrent myocardial infarction, or new-onset heart failure ^38^. Our findings are consistent with multiple previous studies, demonstrating a progressive upward trend in NLR across both ECG and Gensini score groups, with similar proportions of Lym-related inflammatory cells represented by NLR. In ROC curve analysis, the NLR diagnostic AUC for STEMI was 0.838, with a corresponding cutoff value of 2.06, sensitivity of 0.831, and specificity of 0.722. When the proportions of Neu, Mon, and Lym were combined to reflect inflammation, the SIRI AUC was higher at 0.864, with a corresponding cutoff value of 1.03, sensitivity of 0.856, and specificity of 0.738. Our results indicate that the proportion of multiple inflammatory cells is more predictive of disease severity than the proportion of single cells. SIRI integrates three types of inflammatory cells, providing a more accurate reflection of the body’s inflammatory status and immune system function, with superior sensitivity compared to single-component detection. Elevated SIRI values suggest an increase in Neu and/or Mon and/or a decrease in Lym. Multiple studies have confirmed that SIRI levels are associated with prognosis in the general population and patients with cardiovascular diseases ^16,39–41^. Notably, in ROC curve analysis, the sensitivity of SIII and SV was also increased. The AUC for SIII in diagnosing STEMI was 0.815, with a corresponding cutoff value of 22.5, sensitivity of 0.759, and specificity of 0.746. The AUC for SV in diagnosing STEMI was 0.812, with a cutoff value of 8.6, sensitivity of 0.795, and specificity of 0.69. The underlying mechanism of this association may be similar to that of SIRI. Elevated serum levels reflect an imbalance between pro-atherogenic and anti-atherogenic immune networks, which may promote plaque vulnerability, endothelial dysfunction, and thrombotic events, thereby leading to acute coronary events ^42^.Our results demonstrate that SIII and SV, as novel biomarkers, are also important parameters for assessing the severity of ACS patients, and when combined with Eos, these biomarkers may even be more significant than SIRI.

Our results are consistent with previous studies stating that Eos is a strong predictor of cardiovascular disease severity. Therefore, the innovation of our study lies in establishing new inflammatory ratios, SII and SV, which are involved in predicting disease severity, coronary artery stenosis, and cardiac function impairment.

## Limitations

This study has several limitations that need to be considered. First, the data were derived from a single medical center with a relatively small sample size, necessitating larger multicenter studies with extended follow-up periods to validate the findings. Second, confounding factors such as WBC, BMI, age, and medical history were not adequately accounted for. Finally, it would be more valuable if trends in biomarker changes and associations between different outcome indicators could be collected and analyzed over time in patients with varying types of chest pain. The results of this study require further validation in subsequent research.

## Conclusion

This study reveals that EOS is correlated with the severity of ACS, and can predict the severity of ACS as well as the degree of vascular stenosis and the occurrence of heart failure. However, the molecular mechanism underlying its gradual decline remains unclear. The new ratio SIII and SV are also important parameters for assessing the severity of ACS in patients and hold significant clinical importance in emergency evaluation.

## Data Availability

Data is available on request from the first author and corresponding authors.

## Author contributions

C.C., Z.H.Z., L.X., J.N.G., X.L., are involved in the patients’ database analysis, measurement, data analysis and writing. C.C., Z.H.Z.,are involved in statistics, X.Q.X., Y.H.Z., are involved in project design, manuscript review and finalization.

## Ethics Approval declaration

This study was approved by the Science and Ethics Review Committee of the Affiliated Hospital of Yanbian University (Yanbian Hospital) (Approval No.: 20250185) Human Ethics and Consent to Participate declarations: not applicable.

## Clinical trial number

not applicable

---

**Supplementary Figure 1.**
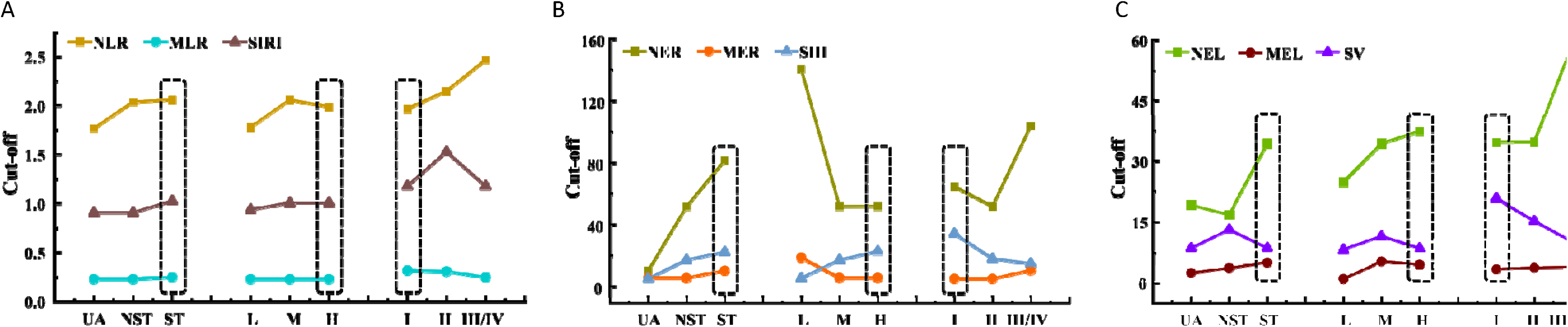
Cut-off values for inflammatory cell ratios in different diagnostic groups of ACS. A. Lym as the denominator for the relative inflammatory ratio in different diagnostic groups of ACS B.Eos as the denominator for the relative inflammatory ratio in different diagnostic groups of ACS C.EosxLym as the denominator for the relative inflammatory ratio in different diagnostic groups of ACS

**Supplementary Table1.**
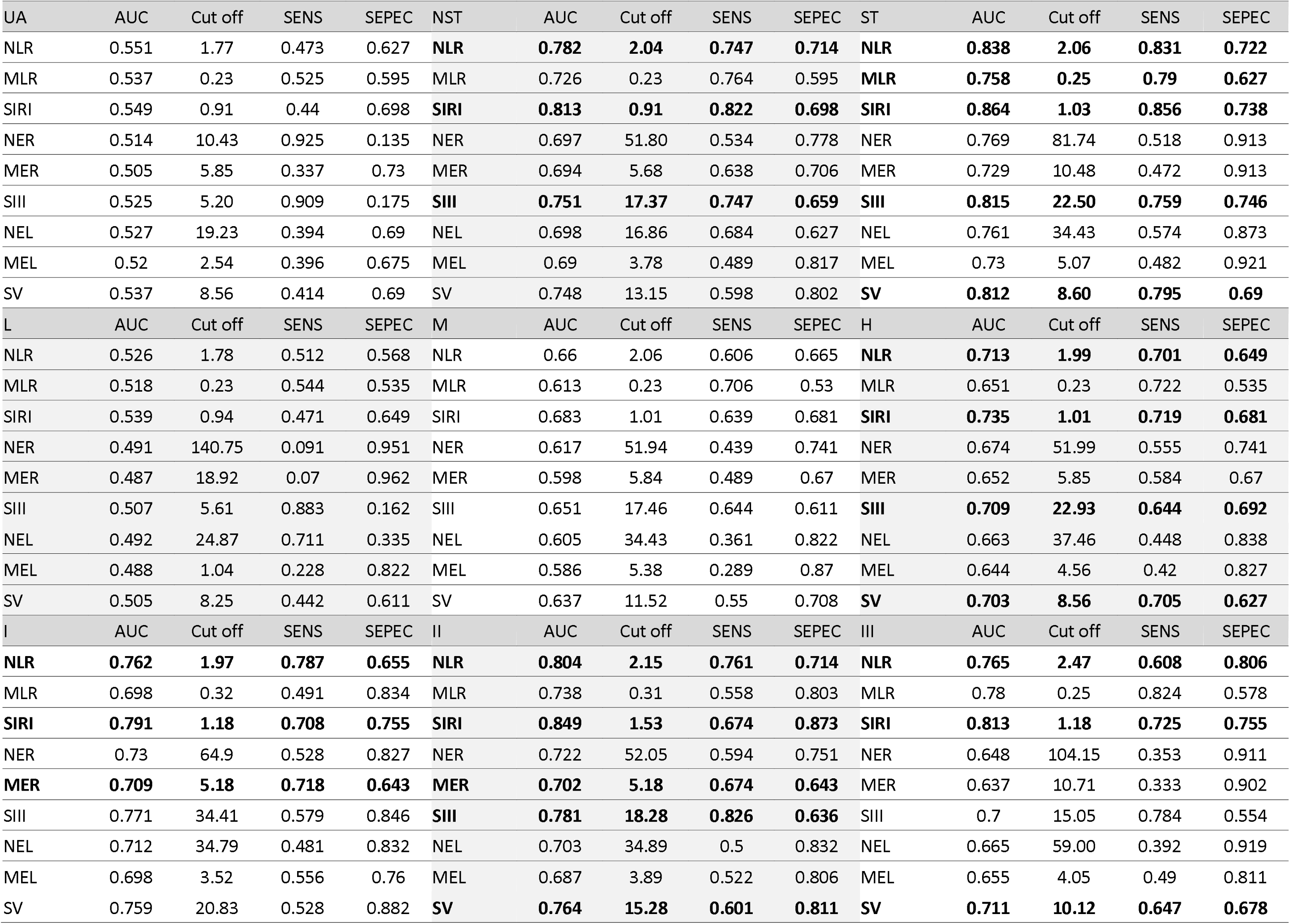
Description of AUC values, cutoff values, sensitivity, and specificity for the proportion of inflammatory cells in each group.

